# Artificial Intelligence Aided Ultrasound Detection of DDH: A Scoping Review Protocol

**DOI:** 10.1101/2024.07.11.24310261

**Authors:** Rusul Yonis, Daniel Perry, James S Bowness, Mohammed Khattak, Natalie Hall

**Affiliations:** Alder Hey NHS Trust, Department of Trauma & Orthopaedics. Liverpool, United Kingdom; Professor of Children’s Orthopedic Surgery & NIHR Research Professor at Alder Hey Children’s, Hospital. Liverpool, United Kingdom; Department of Anesthesia, University College London Hospitals NHS Foundation Trust, London, United Kingdom; Faculty of Health and Life Science, University of Liverpool. Liverpool, United Kingdom; Alder Hey Children’s Hospital. Liverpool, United Kingdom

## Abstract

**Background:** Ultrasound imaging plays a pivotal role in the diagnosis and monitoring of developmental dysplasia of the hip (DDH). However, this step requires a formal referral to the radiology department for an ultrasound by an expert radiologist or sonographer. This process can delay diagnosis and treatment initiation due to long wait times caused by the high demand on NHS services.

In recent years, there has been a growing interest in leveraging artificial intelligence (AI) in ultrasound imaging. AI has potential to assist in image acquisition and interpretation, to inform clinical decision-making. Further benefits may include improved accuracy, efficiency, and consistency in diagnosis, ultimately leading to better patient outcomes.

This scoping review aims to review the evidence for AI to support ultrasound detection of DDH, including reviewing the methodologies employed, the accuracy and utility of algorithms, challenges and opportunities for clinical translation, and requirements for future research.

**Methods:** We will conduct a comprehensive search of the literature using multiple databases, including ACM Digital Library, EMBASE, OVID MEDLINE, PUBMED, COCHRANE Library, CINAHL, and IEEE Explore. These databases cover a wide range of academic disciplines, including computer science, and medical sciences, ensuring thorough coverage of relevant studies related to artificial intelligence (AI) in ultrasound for developmental dysplasia of the hip (DDH).

In addition, we will explore the International Committee of Medical Journal Editors (ICMJE) approved clinical trial registries and the World Health Organization (WHO) clinical trials registry to identify ongoing or completed studies in this field. This will capture relevant research that may not yet be published in peer-reviewed journals.

To supplement the research databases, we will search the websites of international societies in relevant fields, such as the British Society of Children’s Orthopaedic Surgery (BSCOS) and Paediatric Orthopaedic Society of North America (POSNA).

As AI has a strong commercial interest, we will review product information and publicly available evidence from EXO Imaging (https://www.exo.inc), a commercial company with a known interest in this field and an established AI aided US device.

**Discussion:** This scoping review represents the first comprehensive attempt to gather the available evidence on the application of AI in ultrasound imaging for the diagnosis of DDH. By systematically reviewing and synthesizing a diverse range of studies, we aim to provide an overview of the current state of the art in this emerging field, identify gaps in the literature, and inform future research.

## BACKGROUND

Developmental dysplasia of the hip (DDH), is a spectrum of disease ranging from mild dysplasia (i.e., a shallow hip) to a completely dislocated hip.^**(1)**^ It can involve one or both hips. This condition is evident in early infancy and may be influenced by factors such as a genetic predisposition or pressure on the infant in-utero (i.e., a breech position). If left untreated, DDH can result in hip pain, limping, and osteoarthritis later in life.^**(1)**^ Early detection and treatment, often with the use of braces or harnesses to hold the hip joint in place, can help prevent long-term complications.^**(1)**^

The reliance on physical examination for new born screening has been disappointing, as dislocated hips are still diagnosed in later infancy and childhood.^**(1) (2)**^ A recent diagnostic accuracy study demonstrated that if a screening examination was conducted in 1000 hips of newborns, the Barlow/Ortolani manoeuvres would identify 14 dislocations but result in 6 missed diagnoses and 19 unnecessary follow-up diagnostic ultrasound scans. Alternatively, a screening approach using limited hip abduction would identify 2 cases, with 18 missed diagnoses and 32 unnecessary scans amongst 1000 newborn hips.^**(2)**^

Given the poor reliability of clinical screening, ultrasound has become increasingly utilized in diagnosing hip abnormalities.^**(1)**^ The ability of ultrasound to visualize the cartilaginous acetabulum and femoral head makes it an appealing choice for examining the infant hip. In contrast, radiography is not effective in diagnosing this condition at such an early age, as the femoral head and a significant portion of the acetabular roof have yet to ossify. ^**(1)**^ Therefore, ultrasound has emerged as a preferred method for early detection and assessment of developmental dysplasia of the hip. However, the accuracy of the diagnosis depends heavily on expertise of the operator. ^**(1)**^

Recent advancements in medical technology, particularly the emergence of artificial intelligence (AI), have spurred efforts to integrate ultrasound with AI to enhance patient outcomes. This broad term encompasses the ability of computers to perform tasks traditionally associated with human intelligence. In clinical practice, AI-based ultrasound image analyses is gaining traction across a variety of medical fields.^**(3–6)**^ These technologies leverage the power of deep learning algorithms for image analysis to provide valuable insights for healthcare professionals. As AI continues to evolve, its integration into medical imaging holds immense potential for improving diagnostic accuracy, streamlining workflows, and enhancing patient care. Initiatives focus on assisting specialists in promptly diagnosing developmental DDH at an earlier age, thereby reduce the incidence of delayed diagnosis.

Several critical issues remain in implementing and evaluating AI ultrasound systems for DDH. Key challenges include standardizing imaging protocols, validating AI algorithms across diverse populations, and assessing AI’s clinical impact on patient management. This field is inherently interdisciplinary, drawing expertise from computer science, engineering, and clinical medicine. Specialists often diverge in their focus and approaches, leading to potential gaps in communication. Currently, there is a lack of centralised resources that integrate analyses of the diverse range of early studies and identify areas of deficiency/requiring improvement. To address this, a comprehensive review of existing literature and publicly available data is warranted. As of April 16, 2024, a preliminary search did not yield any scoping or systematic reviews registered with the Joanna Briggs Institute (JBI) Database of Systematic Reviews and Implementation Reports, the Cochrane Database of Systematic Reviews, or the International Prospective Register of Systematic Reviews (PROSPERO).

## SCOPING REVIEW OBJECTIVES

We will map the available evidence in this field, identify promising areas for exploration or clinical translation and areas which require further investigation. Ultimately, our goal is to lay the groundwork for continued research and innovation in this rapidly evolving field.

## SCOPING REVIEW PROTOCOL

The protocol presented below was developed using the guidance provided by the JBI and Preferred Reporting Items for Systematic Reviews and Meta-Analyses – Extension for Scoping Reviews (PRISMA-ScR).^**(7,8)**^

## SCOPING REVIEW QUESTION

“What is the evidence supporting the use of artificial intelligence aided ultrasound for the detection of DDH?”

## INCLUSION CRITERIA

### Population

Our search will focus on sources published during or after 1980, which coincides with the introduction of the Graf method for screening infant hip by ultrasound.^**(9)**^ We will only include data from papers centred on detecting DDH in infants less than 6 months of age, the age at which the femoral head ossific nucleus forms rendering ultrasound less effective at scanning hip morphology.^**(10,11)**^

### Concept

The scoping review aims to explore the accuracy of AI-based ultrasound detection of DDH.

### Context

Sources must relate to ultrasound scanning in the context of DDH.

### Types of sources

We will cast a wide net and incorporate diverse sources of information, including peer-reviewed articles, guidelines from reputable medical societies, and promotional materials from commercial entities.

We will not impose restrictions based on the type of ultrasound machine or probe utilized in the studies. Furthermore, we will include all the relevant studies irrespective of the scanning technique used in obtaining the images.

Sources published in languages other than English will be translated and/or the author(s) will be contacted for assistance with data extraction.

## SEARCH STRATEGY

### Published Academic Literature

The search strategy for this scoping review was developed by the lead investigator (RY) and the clinical librarian (NH), then reviewed and modified by the senior team (DP and JSB). The strategy utilised multiple databases, including the ACM Digital Library, EMBASE, OVID MEDLINE, PUBMED, COCHRANE library, CINAHL, and IEEE Explore. Searches will be constrained by date (from 1980 to 2024) and subject matter (limited to human studies, excluding animal research).

During the initial search, keywords outlined in Table 1 will be employed to identify relevant manuscripts.

**Table 1.** Search strategy: The main terms identified to be searched for in title, abstract, author keywords (if available) and subject headings. We will also consider abbreviations and incorporate different combinations of these terms.

| Artificial intelligence | And | Ultrasound | And | Developmental Dysplasia of the Hip |
| --- | --- | --- | --- | --- |
| “Artificial intelligence” |  | “Ultrasound” |  | “DDH*” |
| “Computer vision” |  | “Sonography” |  | “Developmental Dysplasia of the Hip” |
| “Machine intelligence” |  |  |  | “Congenital Hip Dislocation” |
| “Deep learning” |  |  |  | “Hip Dysplasia” |
| “Machine learning” |  |  |  | “Hip Subluxation” |
| “Neural network” |  |  |  | “Hip Dislocation” |
| “Computer aided” |  |  |  |  |
| “AI*” |  |  |  |  |
| “Machine vision” |  |  |  |  |

### Other Literature

We will explore ICMJE and WHO clinical trial registries to identify ongoing or completed studies in this field, capturing research that may not yet be published. In addition to that, we will review websites of relevant international societies, such as BSCOS, and POSNA.

As AI is an area of intense commercial research, other evidence and commercial data will also be sought to minimise publication bias. We will search publicly available information from EXO Imaging (https://www.exo.inc), a large commercial company with a known established commercially available device and interest in this field.

## DATA EXTRACTION

### Screening of title and abstract review

Once all sources have been identified through the search process outlined above, two investigators will independently screen the titles and abstract for inclusion in the study.

If a title is ambiguous and there is uncertainty whether it meets the inclusion criteria, the source will be included in the abstract review stage. In instances where there is disagreement between the assessors, a third investigator will review the title/abstract and provide an adjudication to resolve the disagreement.

### Extraction of Results

The full texts of all relevant sources will undergo comprehensive review to confirm their continued suitability. Results and conclusions will be meticulously examined to extract data regarding the evaluated AI systems. This extracted data will be systematically organized using Table 2 for electronic charting purposes.

**Table 2.**
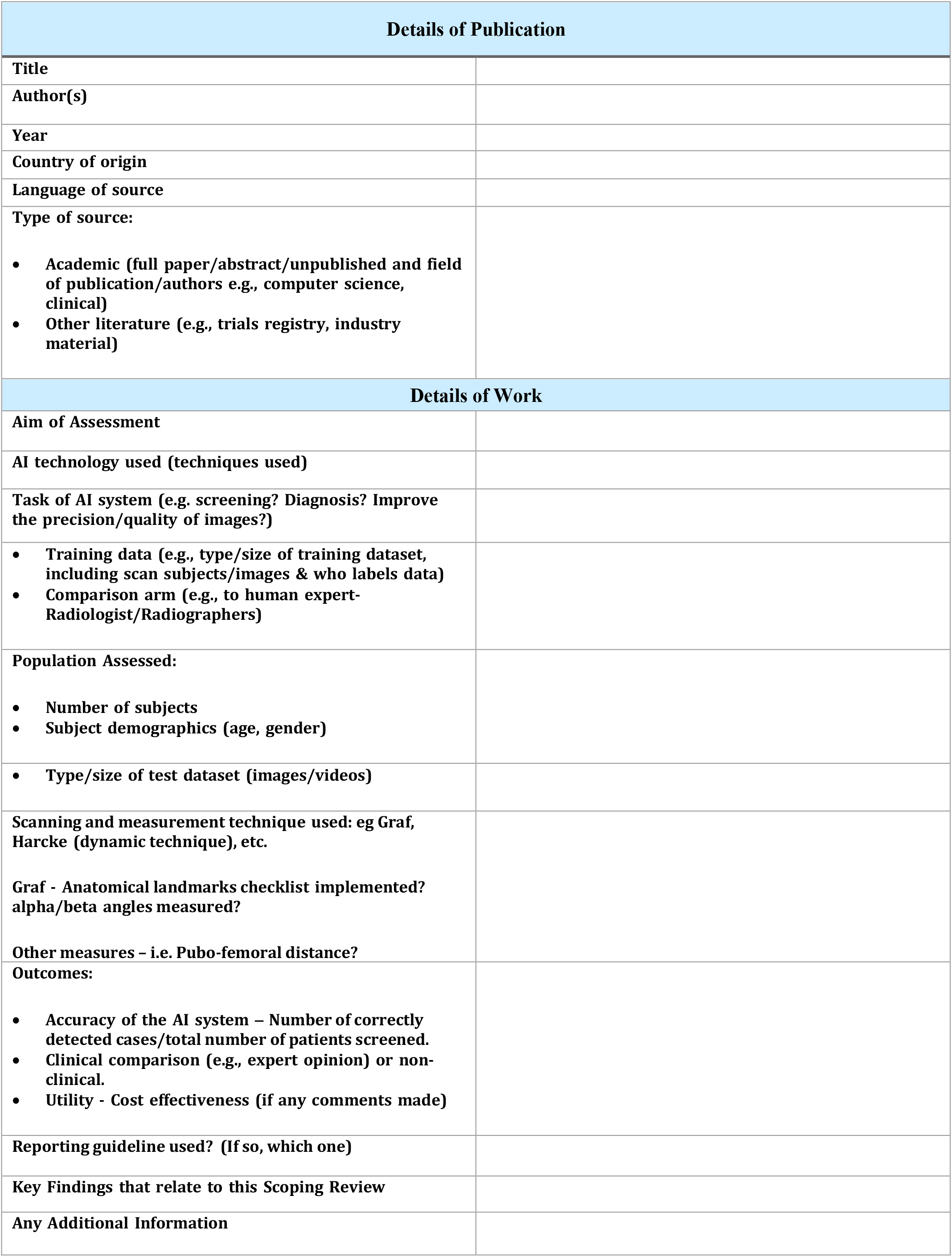
Data Extraction Form.

Given the exploratory nature of this scoping review within an emerging field, we are not setting predefining criteria for minimum outcome/reporting standards. To ensure thorough data capture, an inclusive approach will be employed during the review process. Emphasis will be placed on evaluating aspects such as methods of training/evaluating the AI models, sono-anatomical features of interest used for the models, the ability of AI models to identify DDH, and reporting of the data.

Furthermore, references to ultrasound scanning, whether in a simulation or clinical setting, will be considered relevant as the scanning itself is conducted on human tissue.

## COLLATING AND SUMMARISING DATA, PRESENTATION OF RESULTS

The search and screening process will be visually depicted in a flowchart, outlining the number of publications considered at each stage, from initial search to final inclusion.

The data extracted from this search and review process will be presented in a descriptive format, divided into three main sections:

1. The first will focus on discussing data related to the accuracy of AI systems in detecting DDH on ultrasound. This section will delve into the findings regarding the performance of AI-assisted ultrasound technology in accurately identifying DDH in infants under six months of age.
2. The second will centre on discussing data related standards pertaining to model training (e.g., what ground truth is used for model testing/evaluation), which sono-anatomical structures are used in the model, and consistency/standards of reporting.
3. Lastly, we will discuss this technology’s potential for improving patient outcomes, relating to earlier diagnosis/fewer missed cases and cost-effectiveness of this emerging technology.

## DISCUSSION

The proposed scoping review aims to provide a comprehensive summary of the existing evidence regarding the accuracy and utility of AI systems in ultrasound scanning for the detection of DDH. Given the emerging nature of this field, the quantity and content of available data are expected to vary, potentially reflecting a lack of consistency among studies. Consequently, this review seeks to offer a concise overview of the current knowledge body, identifying key findings and areas which require further investigation. By doing so, we aim to provide valuable guidance for shaping the direction and structure of future research endeavours in this evolving field.

## Data Availability

This is a scoping review to the assess the current knowledge body of the topic addressed.
Will provide the data collected from this scoping review upon reasonable request to the authors.

## LIMITATIONS

Despite the systematic search strategy outlined above, several challenges are anticipated in the proposed study. Firstly, there is a potential for publication bias in the available literature, wherein studies with positive results are more likely to be published than those with negative or inconclusive findings. Furthermore, given the rapid pace of progress in this sector, it is possible that ongoing studies may not yet be publicly available, limiting the comprehensiveness of the review. Moreover, industry data may not be readily accessible, either due to intentional withholding or other reasons, as companies may not always have a commercial incentive to share their data with the public. An additional challenge we anticipate is that some studies may not include patient outcome data, which may hinder the ability to draw conclusive insights regarding the clinical utility of AI systems in ultrasound scanning for DDH detection.

